# Deep learning AI and Restriction Spectrum Imaging for patient-level detection of clinically significant prostate cancer on MRI

**DOI:** 10.1101/2024.11.22.24317504

**Authors:** Yuze Song, Mariluz Rojo Domingo, Christopher C Conlin, Deondre D Do, Madison T Baxter, Anna Dornisch, George Xu, Aditya Bagrodia, Tristan Barrett, Mukesh Harisinghani, Gary Hollenberg, Sophia Kamran, Christopher J Kane, Dimitri A Kessler, Joshua Kuperman, Kanglung Lee, Michael A Liss, Daniel JA Margolis, Paul M Murphy, Nabih Nakrour, Truong Ngyuen, Thomas L Osinski, Rebecca Rakow-penner, Shoumik Roychowdhury, Ahmed S Shabik, Shaun Trecarten, Natasha Wehrli, Eric P Weinberg, Sean A Woolen, Anders M Dale, Tyler M Seibert

## Abstract

**Background:** The Prostate Imaging Reporting & Data System (PI-RADS), based on multiparametric MRI (mpMRI), is widely used for the detection of clinically significant prostate cancer (csPCa, Gleason Grade Group (GG≥2)). However, its diagnostic accuracy can be impacted by variability in interpretation. Restriction Spectrum Imaging (RSI), an advanced diffusion-weighted technique, offers a standardized, quantitative approach for detecting csPCa, potentially enhancing diagnostic consistency and performing comparably to expert-level assessments.

**Purpose:** To evaluate whether combining maximum RSI-derived restriction scores (RSIrs-max) with deep learning (DL) models can enhance patient-level detection of csPCa compared to using PI-RADS or RSIrs-max alone.

**Materials and Methods:** Data from 1,892 patients across seven institutions were analyzed, selected based on MRI results and biopsy-confirmed diagnoses. Two deep learning architectures, 3D-DenseNet and 3D-DenseNet+RSI (incorporating RSIrs-max), were developed and trained using biparametric MRI (bpMRI) and RSI data across two data splits. Model performance was compared using the area under the receiver operating characteristic curve (AUC) for patient-level csPCa detection, using PI-RADS performance for clinical reference.

**Results:** Neither RSIrs-max nor the best DL model combined with RSIrs-max significantly outperformed PI-RADS interpretation by expert radiologists. However, when combined with PI-RADS, both approaches significantly improved patient-level csPCa detection, with AUCs of 0.79 (95% CI: 0.74-0.83; *P*=.005) for combination of RSIrs-max with PI-RADS and 0.81 (95% CI: 0.76-0.85; *P*<.001) for combination of best DL model with PI-RADS, compared to 0.73 (95% CI: 0.68-0.78) for PI-RADS alone.

**Conclusion:** Both RSIrs-max and DL models demonstrate comparable performance to PI-RADS alone. Integrating either model with PI-RADS significantly enhances patient-level detection of csPCa compared to using PI-RADS alone.

**Summary Statement:** RSIrs-max and deep learning models match the performance of expert PI-RADS in patient-level csPCa detection and combining either with PI-RADS yields a significant improvement over PI-RADS alone.

**Key Points:**

- In a study of 1,892 patients from seven institutions undergoing MRI and biopsy for prostate cancer, RSIrs-max and the DL model (AUC, 0.75 (*P*=.59) and 0.78 (*P*=.09)) performed comparably to expert-level PI-RADS scores (AUC, 0.73).
- Including prostate auto-segmentation improved the DL model (AUC, 0.68 (*P*=.01) vs 0.72 (*P*=.60)).
- Combining RSIrs-max or the DL model (AUC, 0.79 (*P*=.005) and 0.81 (*P* <.001)) with PI-RADS statistically significantly outperformed PI-RADS alone (AUC, 0.73).

## Introduction

Multiparametric magnetic resonance imaging (mpMRI) plays a key role in the early diagnosis of prostate cancer, as recommended by the European Association of Urology (EAU) and National Comprehensive Cancer Network (NCCN) guidelines^1^. mpMRI has been shown to reduce unnecessary biopsies and improve the detection of clinically significant prostate cancer (csPCa, grade group (GG)≥2)^1–4^. The Prostate Imaging Reporting & Data System (PI-RADS v2.1) was developed to provide a standardized approach for interpreting mpMRI. However, interpretation can still vary based on the reader’s experience and training. As the incidence of prostate cancer is expected to increase in the coming years, there may be challenges in meeting demand with the current supply of trained experts^6^. An accurate, supportive tool for interpreting prostate MRI could facilitate standardization and address variability in clinical practice^7^.

Restriction Spectrum Imaging (RSI) is an advanced technique for diffusion-weighted imaging (DWI) that measures signal from four distinct tissue compartments: restricted intracellular water (RSI-C_1_), hindered extracellular water (RSI-C_2_), freely diffusing water (RSI-C_3_), and vascular flow (RSI-C_4_)^1,2^. RSI restriction score (RSIrs) is a quantitative biomarker based on RSI that has been shown to be superior to Apparent Diffusion Coefficient (ADC) for detection of csPCa^3–6^. Moreover, when maximum RSIrs (RSIrs-max) is combined with PI-RADS, the performance for patient-level detection of csPCa has been shown to be superior to either alone^4,6^.

PI-RADS relies predominantly on the *T_2_*-weighted imaging (T2w) and DWI components of mpMRI, collectively called biparametric MRI (bpMRI)^7^. There is interest in moving toward bpMRI for many patients, as bpMRI avoids the risks and costs associated with intravenous contrast^8–11^. Deep learning artificial intelligence (AI) models have been developed based on bpMRI for objective and reproducible detection and localization of csPCa, with results matching those of expert radiologists^12–16^.

As both RSI and deep-learning AI models been shown to be accurate and useful for methods, we investigated whether combining the two would improve the automated patient-level detection of csPCa.

## Materials and Methods

### Study Population

The data for this study comes from seven imaging centers participating in the Quantitative Prostate Imaging Consortium (QPIC): the Center for Translational Imaging and Precision Medicine at the University of California San Diego (CTIPM), UC San Diego Health (UCSD), University of California San Francisco (UCSF), Harvard University affiliated Massachusetts General Hospital (MGH), University of Rochester Medical Center (URMC), University of Texas Health Sciences Center San Antonio (UTHSCSA), and University of Cambridge (Cambridge)^6^. The study was approved by each center’s institutional review board (IRB). At UTHSCSA and Cambridge, the data were collected prospectively as part of related projects with written informed consent. At the other centers, the data were collected retrospectively, and a waiver of consent was approved by the respective IRBs for secondary use of routine clinical data. Individuals were included if aged ≥18 years and underwent prostate MRI for suspected PCa or active surveillance between January 2016 and March 2024. They were excluded if they had prior treatment of PCa or if there was no available biopsy result from within 6 months of a positive MRI scan (PI-RADS≥3). Patients with metallic implants were also excluded to avoid metal-induced imaging artifacts. The diagnosis of csPCa was confirmed on biopsy histopathology as per standard-of-care practice at each center. These data have been previously analyzed for performance of RSIrs-max^4,6,17^ (Figure 1).

**Figure 1.**
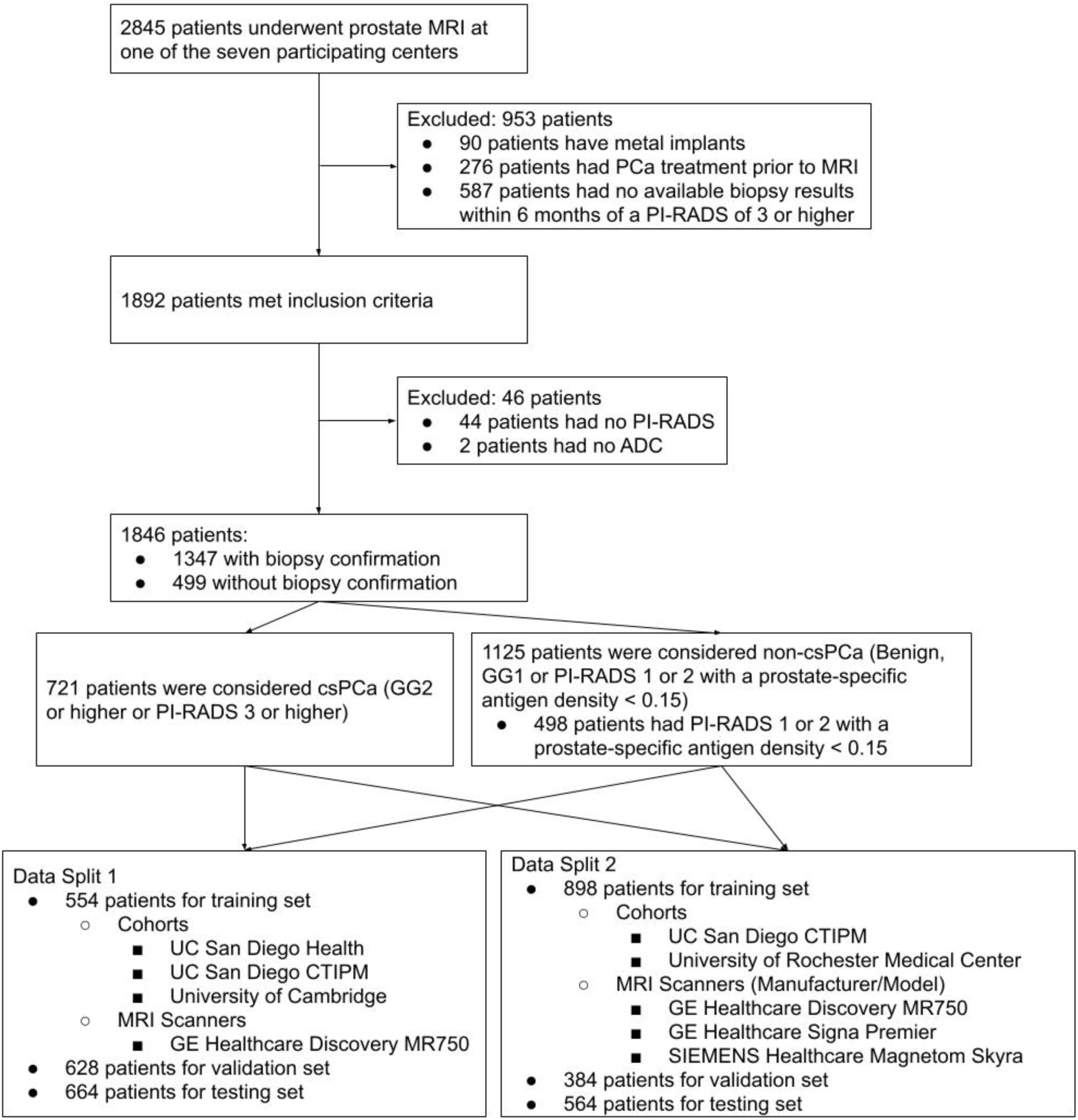
Flowchart shows inclusion and exclusion criteria and patient characteristics. PCa = Prostate Cancer. PI-RADS = Prostate Imaging Reporting and Data System. csPCa = Clinically Significant Prostate Cancer. ADC = Apparent Diffusion Coefficient. GG = Gleason Grade Group. UC = University of California. CTIPM = Center for Translational Imaging and Precision Medicine

### Data acquisition and processing

The processing for RSI data included correction for background noise, eddy currents, and gradient nonlinearities^18–20^. Correction for distortion caused by ***B_0_*** inhomogeneity was applied to data acquired at CTIPM^21^. ADC, DWI and RSI data were resampled to the same image resolution as the T2w data. Automated prostate contours were obtained using an FDA-cleared commercial product (OnQ Prostate, CorTechs.ai, San Diego, CA).

For RSI data, the signal intensity for each b-value was modeled as a linear combination of exponential decays representing four diffusion compartments (RSI-C_1_, RSI-C_2_, RSI-C_3_ and RSI-C_4_), each with a diffusion coefficient determined empirically in previous work^2^. The RSIrs biomarker is the intensity value of the RSI-C_1_ signal at a given voxel normalized by median *T_2_*-weighted signal in the prostate. RSIrs-max is the maximum RSIrs value within a given patient’s prostate. Additional details regarding the RSI modeling are provided in Supplementary Materials.

### Model

RSIrs-max-only and PI-RADS-only were previously analyzed for performance of patient-level csPCa detection using univariable logistic regression^6^. Here, we compare the performance of deep learning models to those previously described logistic regression models. 3D-DenseNet^21^ and 3D-DenseNet+RSI, both 3D densely connected convolutional networks, were trained to get the probability of csPCa with different modalities. The details of the models are illustrated in Figure 2.

**Figure 2.**
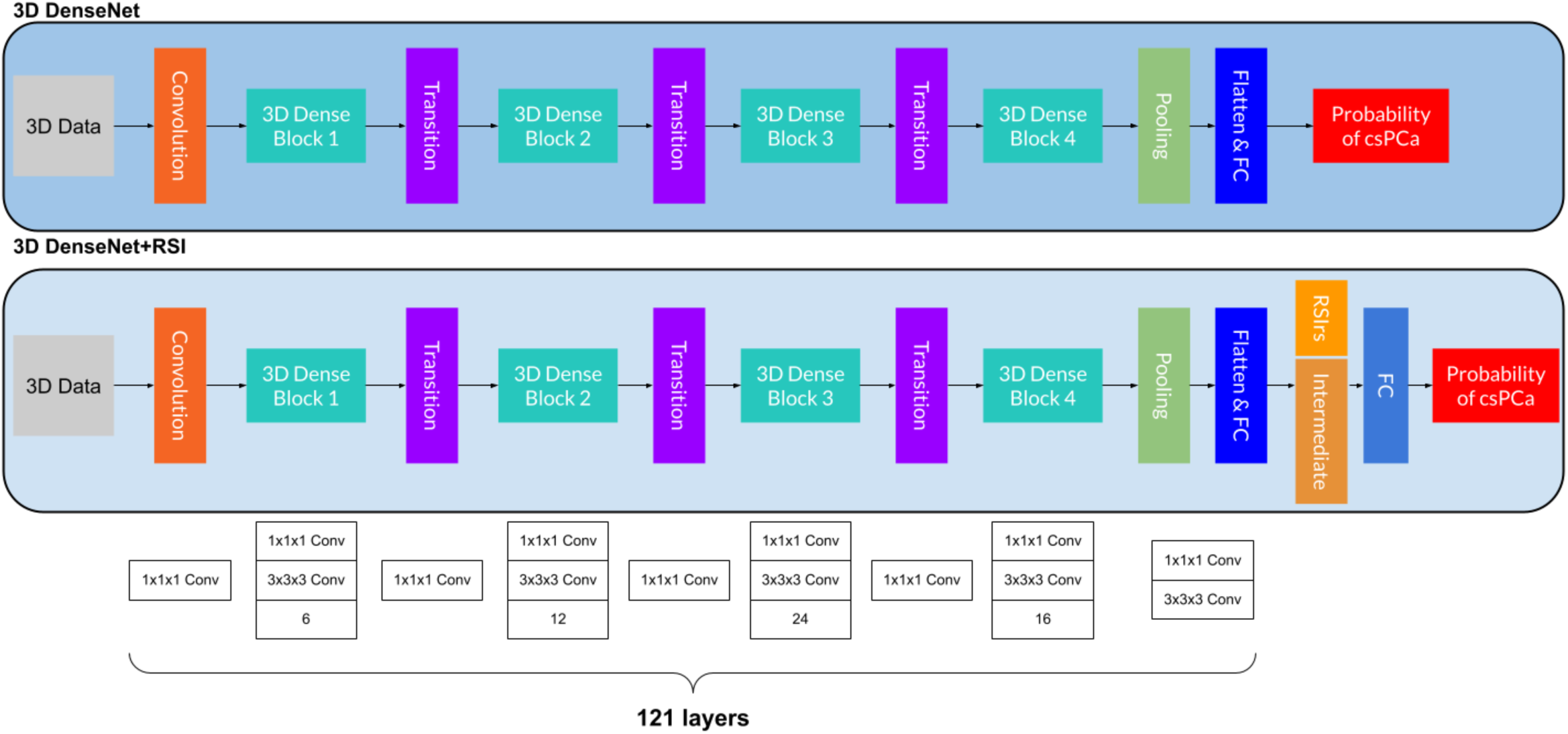
Overview of the experimental setup. 3D-Data are used as input of the 3D-DenseNet, followed by convolution layer, several 3D Dense Blocks and Transition layers, then after the Pooling layer and Flatten & Fully Connected (FC) layer, the 3D-DenseNet returns the final classification of csPCa or not. For 3D-DenseNet+RSI, the process before the Flatten & FC layer is the same, the remaining process is concatenating the intermediate output from the Flatten & FC layer with the RSIrs-max. Then, after going through another FC layer, the 3D-DenseNet+RSI gives the final output of probability of csPCa.

The loss function used to supervise the two models is the Cross Entropy Loss (*Loss*_*CE*_):

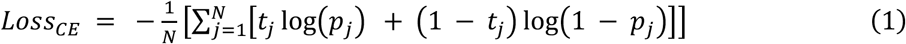

In the equation (1), *N* is the number of data points. *t_j_* refers to the ground truth value. If the patient has a csPCa, then *t_j_* is 1. *p_j_* is the SoftMax probability for the *i^*t*h^* data point.

We also use either RSIrs-max or output probabilities of the AI models, along with PI-RADS, as inputs to a multivariable logistic regression model for comparison with either alone.

### Implementation Details

We employed two different data splits for this study (Figure 1). Data Split 1 was used to test all models and was chosen to facilitate direct comparison with a previous study^6^, which also used Data Split 1; Data Split 2 allocated a larger and more diverse dataset for training because deep learning models’ performance typically improves substantially when trained on heterogeneous data. Data Split 2 was used for testing the model with RSIrs-max-only input; the PI-RADS-only input; the best-performing deep learning model; the combination of RSIrs-max and PI-RADS; and the combination of the output probability from the best-performing deep learning model and PI-RADS from analysis of Data Split 1. We also divided and tested the data for both splits based on GG.

Data Split 1 was described previously^6,17^. Scans collected with the RSI acquisition protocol and the same scanner model (GE Healthcare Discovery MR750) were used for training (n=554). The remaining protocols were used for the testing dataset (n=664) with patients who were biopsy-naïve at the time of the MRI scan and received a biopsy following the MR acquisition. The validation dataset (n=628) is from the rest of the patients that are not included in either training or testing data of Data Split 1 (Table 1).

**Table 1.** Patient Demographic Characteristics in the Data Sets from Data Split 1 and Data Split 2. Unless otherwise specified, data are numbers of examinations, with percentages in parentheses. *The training set for Data Split 1 were acquired with the same RSI protocol. **Data are median with IQR in parentheses. csPCa = Clinically Significant Cancer (Gleason Grade Group ≥ 2 or PI-RADS ≥ 3). non-csPCa = Not having Clinically Significant Cancer (Benign, Gleason Grade Group = 1 or PI-RADS < 3 with a prostate-specific antigen density < 0.15). CTIPM = Center for Translational Imaging and Precision Medicine. UC = University of California. UT = University of Texas. PI-RADS = Prostate Imaging Reporting & Data System.

| Characteristic | Training (Data Split 1)* | Validation (Data Split 1) | Testing (Data Split 1) | Training (Data Split 2) | Validation (Data Split 2) | Testing (Data Split 2) |
| --- | --- | --- | --- | --- | --- | --- |
| No. of cases | 554 | 628 | 664 | 898 | 564 | 384 |
| Age (y)** | 70 (64-75) | 70 (65-75) | 69 (64-74) | 70 (64-74) | 70 (65-75) | 70 (64-75) |
| csPCa | 192 (34.7) | 184 (29.3) | 345 (52.0) | 376 (41.9) | 116 (20.6) | 229 (59.6) |
| non-csPCa | 362 (65.3) | 444 (70.7) | 319 (48.0) | 522 (58.1) | 448 (79.4) | 155 (40.4) |
| Cohorts |  |  |  |  |  |  |
| UC San Diego Health | 546 (98.6) | 48 (7.7) | 98 (14.8) | 0 (0.0) | 383 (67.9) | 309 (80.5) |
| UC San Diego CTIPM | 3 (0.5) | 386 (61.4) | 258 (38.9) | 647 (72.0) | 0 (0.0) | 0 (0.0) |
| Harvard University's Massachusetts General Hospital (MGH) | 0 (0.0) | 25 (4.0) | 38 (5.7) | 0 (0.0) | 25 (4.4) | 38 (9.9) |
| University of Rochester Medical Center (URMC) | 0 (0.0) | 18 (2.9) | 233 (35.1) | 251 (28.0) | 0 (0.0) | 0 (0.0) |
| UC San Francisco (UCSF) | 0 (0.0) | 22 (3.5) | 20 (3.0) | 0 (0.0) | 22 (3.9) | 20 (5.2) |
| UT Health Sciences Center San Antonio (UTHSCSA) | 0 (0.0) | 129 (20.5) | 17 (2.5) | 0 (0.0) | 129 (22.9) | 17 (4.4) |
| University of Cambridge | 5 (0.9) | 0 (0.0) | 0 (0.0) | 0 (0.0) | 5 (0.9) | 0 (0.0) |
| Manufacturer/Model |  |  |  |  |  |  |
| GE Healthcare/Discovery MR750 | 554 (100.0) | 278 (44.3) | 189 (28.5) | 470 (52.3) | 340 (60.3) | 211 (55.0) |
| GE Healthcare/Signa Premier | 0 (0.0) | 203 (32.3) | 225 (33.9) | 177 (19.7) | 95 (16.8) | 156 (40.6) |
| SIEMENS Healthcare/Magnetom Skyra | 0 (0.0) | 59 (9.4) | 250 (37.6) | 251 (28.0) | 41 (7.3) | 17 (4.4) |
| SIEMENS Healthcare/Magnetom Trio | 0 (0.0) | 88 (14.0) | 0 (0.0) | 0 (0.0) | 88 (15.6) | 0 (0.0) |
| Biopsy |  |  |  |  |  |  |
| Biopsy-confirmed cases | 322 (58.1) | 361 (57.5) | 664 (100.0) | 689 (76.7) | 274 (48.6) | 384 (100.0) |
| Biopsy-naïve cases | 400 (72.2) | 164 (26.1) | 664 (100.0) | 613 (68.3) | 231 (41.0) | 384 (100.0) |
| Gleason Grade Group |  |  |  |  |  |  |
| Benign | 72 (13.0) | 80 (12.7) | 179 (27.0) | 186 (20.7) | 66 (11.7) | 79 (20.6) |
| 1 | 58 (10.5) | 97 (15.5) | 140 (21.1) | 127 (14.1) | 92 (16.3) | 76 (19.8) |
| 2 | 85 (15.3) | 88 (14.0) | 181 (27.3) | 189 (21.0) | 65 (11.5) | 100 (26.0) |
| 3 | 56 (10.1) | 54 (8.6) | 90 (13.6) | 104 (11.6) | 23 (4.1) | 73 (19.0) |
| 4-5 | 51 (9.2%) | 42 (6.7) | 74 (11.0) | 83 (9.3) | 28 (5.0) | 56 (14.6) |
| PI-RADS score |  |  |  |  |  |  |
| 1 | 255 (46.0) | 320 (51.0) | 60 (9.0) | 261 (29.1) | 342 (60.6) | 32 (8.3) |
| 2 | 17 (3.1) | 24 (3.8) | 12 (1.8) | 29 (3.2) | 22 (3.9) | 2 (0.5) |
| 3 | 46 (8.3) | 62 (9.9) | 155 (23.3) | 157 (17.5) | 52 (8.2) | 54 (14.1) |
| 4 | 120 (21.7) | 118 (18.8) | 215 (32.4) | 208 (23.1) | 88 (15.6) | 157 (40.9) |
| 5 | 116 (20.9) | 104 (16.5) | 222 (33.5) | 243 (27.1) | 60 (10.7) | 139 (36.2) |

The training set for Data Split 2 (n=898) includes data from different RSI protocols and collected from two centers (UCSD CTIPM and URMC). Data from UCSD CTIPM were collected using GE Healthcare Discovery MR750 and GE Healthcare Signa Premier scanners. Data from URMC were collected from SIEMENS Magnetom Skyra scanners. The testing dataset (n=384) is from the remaining patients of all other cohorts who were biopsy-naïve at the time of the MRI scan and received a biopsy following the MR acquisition. The validation dataset (n=564) for Data Split 2 is from the rest of the patients that are not included in either training or testing data of Data Split 2 (Table 1). Both testing and validation datasets are external to the training dataset.

Two univariable logistic regression models (RSIrs-max or PI-RADS as input, respectively) and two multi-variable logistic regression models (combination of RSIrs-max or output probability of the best performing AI model with PI-RADS as inputs, respectively) were implemented using MATLAB.

The RSI-C_1_ and RSI-C_2_ components from RSI data, T2w, ADC and high b-value DWI (high-b DWI) were included as the 3D-data for the input of the corresponding model. The preprocessing and augmentation of the input image data is described in Supplementary Materials. The following models were evaluated: Model **1**: PI-RADS; Model **2**: RSIrs-max; Model **3** (bpMRI): T2w, ADC, high-b DWI; Model **4** (bpMRI-seg): T2w with automated prostate segmentation applied as a binary mask (T2w-seg), ADC-seg, high-b DWI-seg; Model **5**: bpMRI-seg, RSIrs-max; Model **6** (bpMRI-seg, RSI-seg, RSIrs-max): bpMRI-seg, RSI-C_1_-seg, RSI-C_2_-seg, RSIrs-max; Model **7**: RSIrs-max, PI-RADS and Model **8**: bpMRI-seg, RSI-seg, RSIrs-max, PI-RADS (Table 2 and Table 3). For all inputs labeled with “-seg”, the automated prostate segmentation was applied to the 3D data as a binary mask through element-wise multiplication.

**Table 2.**
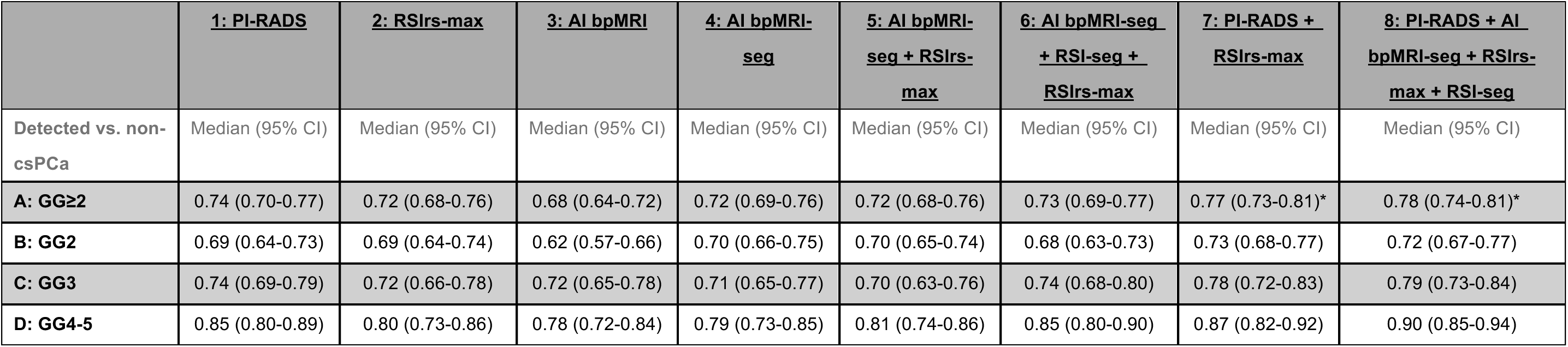
AUCs Results for Data Split 1. 95% CI refers to the 95 percent confidence interval. * refers to *p*<0.05 for comparison of AUC vs. PI-RADS 1 (only reported for primary analysis, GG≥2). Model **1** refers to the logistic regression model with PI-RADS input; Model **2** refers to the logistic regression model with RSIrs-max input; Model **3** refers to the 3D DenseNet with T2w, ADC and high-b DWI input (bpMRI); Model **4** refers to the the 3D-DenseNet with T2w-seg, ADC-seg and high-b DWI-seg input (bpMRI-seg); Model **5** refers to the 3D-DenseNet+RSI with bpMRI-seg and RSIrs-max input; Model **6** refers to the 3D-DenseNet+RSI with bpMRI-seg, RSI-C_1_-seg, RSI-C_2_-seg (RSI-seg) and RSIrs-max input; Model **7** refers to the logistic regression model with PI-RADS and RSIrs-max input; Model **8** refers to the logistic regression model with PI-RADS and the output probability of Model **6**. Group A) All the patients that are biopsy-naïve at time of MRI with biopsy confirmed diagnosis that are not used for training vs. non-csPCa (n=664). Group B) Subsets of Group A) with either GG2 csPCa and non-csPCa (n=500). Group C) Subsets of Group A) with either GG3 csPCa and non-csPCa (n=409). Group D) Subsets of Group A) with either GG4-5 csPCa and non-csPCa (n=393). n is the number of cases in each testing group.

|  | **1: PI-RADS** | **2: RSIRS-max** | **3: AI bpMRI** | **4: AI bpMRI-seg** | **5: AI bpMRI-seg + RSIRS-max** | **6: AI bpMRI-seg + RSI-seg + RSIRS-max** | **7: PI-RADS + RSIRS-max** | **8: PI-RADS + AI bpMRI-seg + RSIRS-max + RSI-seg** |
| --- | --- | --- | --- | --- | --- | --- | --- | --- |
| **Detected vs. non-csPCa** | Median (95% CI) | Median (95% CI) | Median (95% CI) | Median (95% CI) | Median (95% CI) | Median (95% CI) | Median (95% CI) | Median (95% CI) |
| **A: GG≥2** | 0.74 (0.70-0.77) | 0.72 (0.68-0.76) | 0.68 (0.64-0.72) | 0.72 (0.69-0.76) | 0.72 (0.68-0.76) | 0.73 (0.69-0.77) | 0.77 (0.73-0.81)* | 0.78 (0.74-0.81)* |
| **B: GG2** | 0.69 (0.64-0.73) | 0.69 (0.64-0.74) | 0.62 (0.57-0.66) | 0.70 (0.66-0.75) | 0.70 (0.65-0.74) | 0.68 (0.63-0.73) | 0.73 (0.68-0.77) | 0.72 (0.67-0.77) |
| **C: GG3** | 0.74 (0.69-0.79) | 0.72 (0.66-0.78) | 0.72 (0.65-0.78) | 0.71 (0.65-0.77) | 0.70 (0.63-0.76) | 0.74 (0.68-0.80) | 0.78 (0.72-0.83) | 0.79 (0.73-0.84) |
| **D: GG4-5** | 0.85 (0.80-0.89) | 0.80 (0.73-0.86) | 0.78 (0.72-0.84) | 0.79 (0.73-0.85) | 0.81 (0.74-0.86) | 0.85 (0.80-0.90) | 0.87 (0.82-0.92) | 0.90 (0.85-0.94) |

**Table 3.** AUCs Results for Data Split 2. Same as the setting of Table 2. 95% CI refers to the 95 percent confidence interval. * refers to *P*<.05 for comparison of AUC vs. PI-RADS 1 (only reported for primary analysis, GG≥2). Model **1** refers to the logistic regression model with PI-RADS input; Model **2** refers to the logistic regression model with RSIrs-max input; Model **6** refers to the 3D-DenseNet+RSI with bpMRI-seg, RSI-seg and RSIrs-max input; Model **7** refers to the logistic regression model with PI-RADS and RSIrs-max input; Model **8** refers to the logistic regression model with PI-RADS and the output probability of Model **6**. Group E) All the patients that are biopsy-naïve at time of MRI with biopsy confirmed diagnosis that are not used for training vs. non-csPCa (n=384). Group F) Subsets of Group E) with either GG2 csPCa and non-csPCa (n=255). Group G) is subset of Group E) with either GG3 csPCa and non-csPCa (n=228). Group H) is subset of Group E) with either GG4-5 csPCa and non-csPCa (n=211). n is the number of cases in each testing group.

|  | **<u>1: PI-RADS</u>** | **<u>2: RSIRS-max</u>** | **<u>6: AI bpMRI-seg + RSI-seg+ RSIRS-max</u>** | **<u>7: PI-RADS + RSIRS-max</u>** | **<u>8: PI-RADS + AI bpMRI-seg + RSIRS-max + RSI-seg</u>** |
| --- | --- | --- | --- | --- | --- |
| **Detected vs. non-csPCa** | Median (95% CI) | Median (95% CI) | Median (95% CI) | Median (95% CI) | Median (95% CI) |
| **A: GG≥2** | 0.73 (0.68-0.78) | 0.75 (0.70-0.80) | 0.78 (0.73-0.83) | 0.79 (0.74-0.83)* | 0.81 (0.76-0.85)* |
| **B: GG2** | 0.69 (0.63-0.75) | 0.72 (0.66-0.75) | 0.73 (0.66-0.79) | 0.75 (0.69-0.81) | 0.76 (0.70-0.82) |
| **C: GG3** | 0.74 (0.67-0.80) | 0.75 (0.68-0.82) | 0.8 (0.73-0.86) | 0.8 (0.73-0.86) | 0.82 (0.75-0.88) |
| **D: GG4-5** | 0.80 (0.74-0.86) | 0.80 (0.72-0.87) | 0.86 (0.80-0.92) | 0.85 (0.79-0.91) | 0.89 (0.83-0.94) |

All models, preprocessing, and data augmentation were implemented using the PyTorch toolbox and MONAI^22^. The training details is described in Supplementary Materials.

### Statistical analysis

The precise location and extent of csPCa in each patient’s prostate are generally unknown, and targeted biopsy can have MRI-to-ultrasound registration or needle placement errors. Many cancers are also detected through systematic biopsy. We therefore focus on patient-level csPCa detection, a more reliable approach that addresses the key clinical question of whether to recommend an invasive biopsy. The performance for patient-level detection was assessed by the Area Under the Curve of the Receiver Operating Characteristic curve (AUC). For the two data splits, we made statistical comparisons via 10,000-bootstrapping samples to calculate 95% confidence intervals and *P* for the difference between the performance of the models and PI-RADS^23^. The primary analysis was for detection of GG≥2 (csPCa) versus GG=1 and Benign (non-csPCa), for which we determined the statistical significance through a two-sided α=0.05. Secondary subgroup analyses evaluated the results per GG. For Data Split 2, we repeated the above analyses for PI-RADS, RSIrs-max, and the best performing deep learning model (based on median AUC for GG≥2) from the Data Split 1 analyses.

We trained a model with bpMRI and compared its performance with PI-RADS-only and RSIrs-max-only univariable logistic regression models. To assess whether automated prostate contours could enhance performance, we also tested our model with bpMRI-seg input. Finally, we evaluated combining RSIrs-max with bpMRI-seg to determine if RSIrs-max improves performance over bpMRI-seg alone.

Codes used in developing the DL model are available on GitHub.

## Results

Data were acquired using 7 distinct acquisition protocols, 2 scanner vendors, 4 scanner models, and 17 MRI scanners (Table 1, Supplementary Table 2 and 3). 1,892 patients met the inclusion criteria (Figure 1). Occlusion sensitivity map^24^ was generated for interpretation of the DL models (Figure 3 and 4).

**Figure 3.**
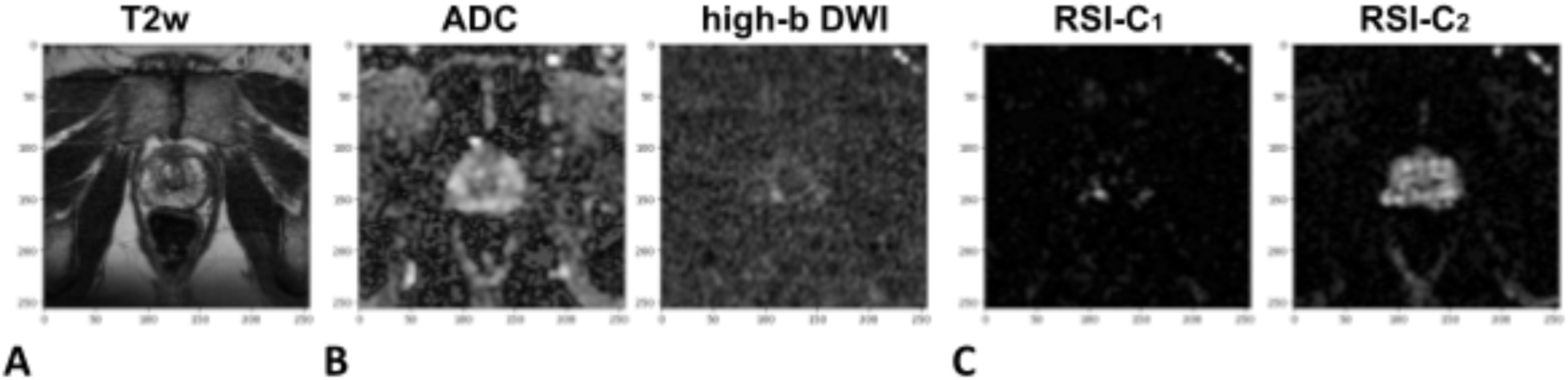
Male patient, age between 55 and 59, underwent MRI due to clinical suspicion of prostate cancer. The patient subsequently underwent prostatectomy and had Gleason Score 4 + 3 (GG 3) cancer at the right and left posterior near the apex of the prostate. He was included in both Data Split 1 and Data Split 2 test sets. The patient-level probability from Model **4** (AI bpMRI-seg) with Data Split 1 was 0.18, from Model **6** (AI bpMRI-seg + RSI-seg + RSIrs-max) with Data Split 1 was 0.47, and from Model **6** with Data Split 2 was 0.51. The radiologists graded this examination as PI-RADS 4 for the lesion at the right peripheral zone at the posterior medial prostate within apex. **(A)** *T_2_*-weighted image (T2w, representative slice). **(B)** Apparent diffusion coefficient map (ADC, representative slice, left) and high-b diffusion-weighted image (high-b DWI, representative slice, right). **(C)** Intracellular (RSI-C_1_, representative slice, left) and extracellular (RSI-C_2_, representative slice, right) images.

**Figure 4.**
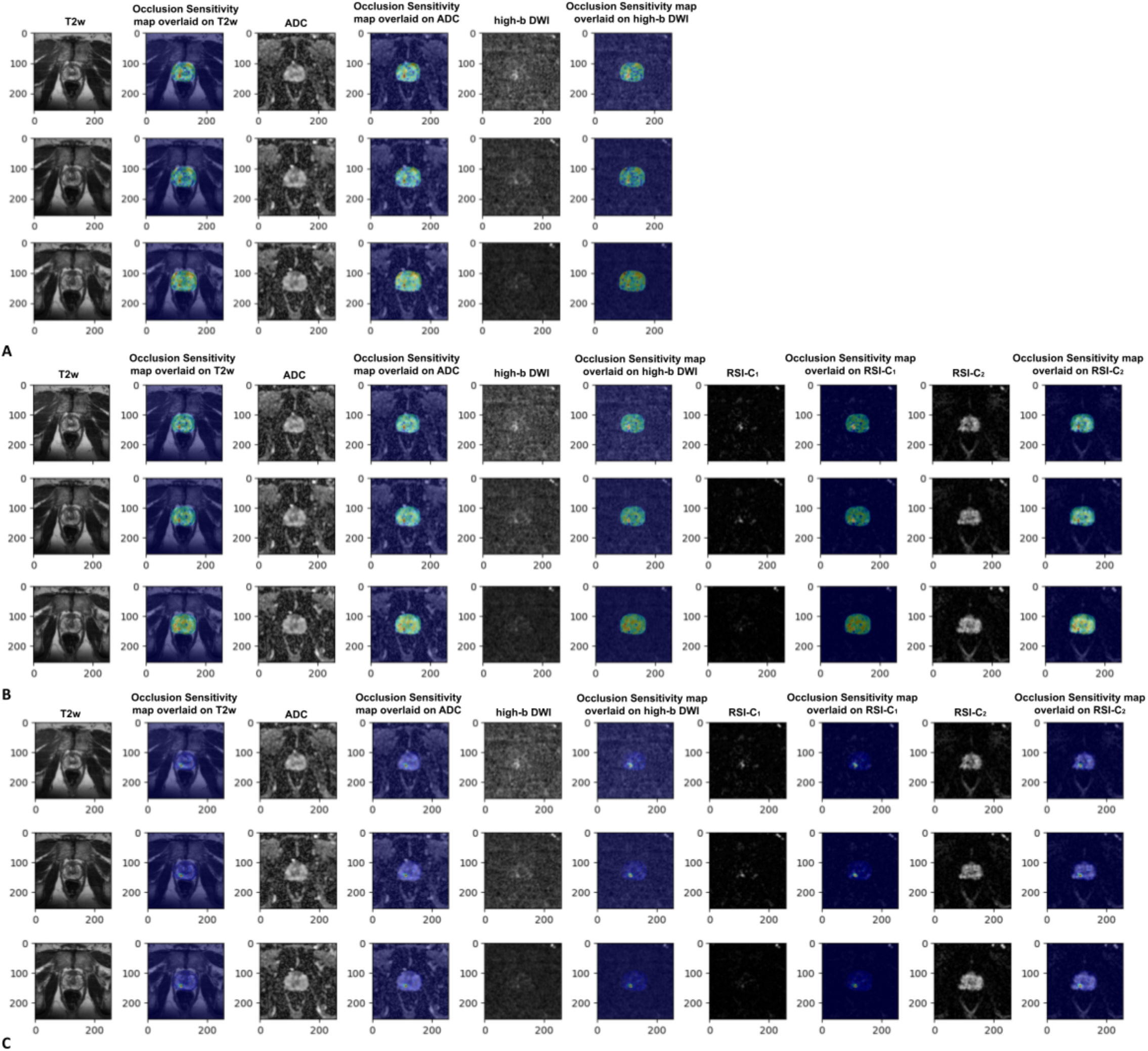
Scan from the same patient as Figure 3. **(A)** T2w, ADC, and high-b DWI images from left to right and with occlusion sensitivity maps from Model **4** (Data Split 1) overlaid at the right of each MRI images. **(B)** T2, ADC, high-b DWI, RSI-C_1_, and RSI-C_2_ images from left to right and with occlusion sensitivity maps from Model **6** (Data Split 1) overlaid at the right of each MRI images. **(C)** T2w, ADC, high-b DWI, RSI-C_1_, and RSI-C_2_ images from left to right with occlusion sensitivity maps from Model **6** (Data Split 2) overlaid at the right of each MRI images.

### Patient-level detection of csPCa for Data Split 1

Table 2 shows the AUC values of models with different 3D-inputs and testing groups for Data Split 1. We tested our model on 4 groups, none of which were used during training. Group A (n=664) consists of patients that were biopsy-naïve at time of MRI and then either had a biopsy to determine cPCa status or were presumed non-csPCa based on low clinical suspicion. Group B (n=500) is the subset of Group A consisting of patients with GG2 csPCa or non-csPCa (i.e., excluding GG≥3). Group C (n=409) is the subset of Group A consisting of patients with GG3 csPCa or non-csPCa. Lastly, Group D (n=393) is the subset of Group A consisting of patients with high grade GG4-5 csPCa or non-csPCa (Table 1).

When an automated prostate segmentation was included (AUC, 0.72 (95% CI: 0.69-0.76; *P*=.60)), patient-level csPCa detection was comparable to PI-RADS (AUC, 0.74 (95% CI: 0.70-0.77)), whereas training the bpMRI model without the prostate segmentation (AUC, 0.68 (95% CI: 0.64-0.72; *P*=.01)) yielded worse performance than PI-RADS. The same pattern was observed for the subgroups based on GG.

Combining PI-RADS with either RSIrs-max (AUC, 0.77 (95% CI: 0.73-0.81; *P*=.01)) or with the best deep learning model (Model **6**) (AUC, 0.78 (95% CI: 0.74-0.81; *P*<.001)) yielded significantly improved patient-level csPCa detection compared to PI-RADS alone (AUC, 0.74 (95% CI: 0.70-0.77)).

The quantitative biomarker RSIrs-max performed comparably in comparison of PI-RADS (*P*=.49) and the bpMRI-seg deep learning AI model (*P*=.85). Combining RSIrs-max with the deep learning bpMRI-seg model did not result in a statistically significant improvement in csPCa detection with PI-RADS, with (*P*=.87) or without (*P*=.49) the 3D volumes from RSI-C_1_ and RSI-C_2_. This suggests that a straightforward, interpretable biomarker (RSIrs-max) derived from approximately two minutes of RSI acquisition may capture most of the valuable information provided by bpMRI. However, when PI-RADS was combined with RSIrs-max (*P*=.01), performance was significantly superior to PI-RADS alone. Likewise, combining PI-RADS with the best deep learning model (*P*<.001) also improved performance beyond PI-RADS alone.

### Patient-level detection of csPCa for Data Split 2

The best-performing AI model in Data Split 1 analyses (based on median AUC) was Model **6**. We therefore calculated and compared AUC values of Models **1**, **2** and **6** within Data Split 2 (Table 3). Group E (n=384) is the full testing dataset for Data Split 2, consisting of either the patients that are biopsy-naïve at time of MRI with a biopsy confirmed diagnosis or are non-csPCa. Group F (n=255) is a subset of Group E consisting of patients with GG2 csPCa and non-csPCa. Group G (n=228) is another subset of Group E consisting of patients with GG3 csPCa and non-csPCa. Lastly, Group H (n=211) is also another subset of Group E consisting of patients with GG4-5 csPCa and non-csPCa.

RSIrs-max (AUC, 0.75 (95% CI: 0.70-0.80; *P*=.59)) and the AI model (Model **6**) (AUC, 0.78 (95% CI: 0.73-0.83; *P*=.09)) had higher point estimates for AUC than PI-RADS (AUC, 0.73 (95% CI: 0.68-0.78)) in Data Split 2, but there was no statistically significant difference between any of these models. Interestingly, though, while the AUC for PI-RADS was similar in the two data splits, the AI model performed better when trained on more diverse data: (AUC, 0.73 (95% CI: 0.69-0.77)) in the testing set for Data Split 1 and (AUC, 0.78 (95% CI: 0.73-0.83)) in the testing set for Data Split 2. Importantly, combining PI-RADS with either RSIrs-max (AUC, 0.79 (95% CI: 0.74-0.83; *P*=.005)) or AI model (AUC, 0.81 (95% CI: 0.76-0.85; *P*<.001)) significantly outperformed PI-RADS alone.

## Discussion

Previous work demonstrated the promising utility of RSIrs-max as a quantitative imaging biomarker for csPCa^2–4,6,25^. In this study, we explored deep learning AI as an alternative or complementary approach for reproducible, objective interpretation of prostate MRI. We observed comparable performance in csPCa detection using PI-RADS, a bpMRI AI model, and RSIrs-max. While training AI models with bpMRI and RSI data (RSIrs-max alone or with full RSI volumes) showed a numerical improvement in detection, it was not statistically significant. However, combining PI-RADS with either the best AI model (Model **6**) or RSIrs-max significantly enhanced detection performance compared to PI-RADS alone. Thus, RSIrs-max provides rapid, quantitative, standardized data to support radiologist interpretation and could augment PI-RADS scoring.

An advantage of a bpMRI AI tool over RSIrs-max is that it may be applicable to datasets lacking DWI compatible with calculation of RSIrs-max. The deep learning model does not rely on the radiologist’s expertise and offers results comparable to PI-RADS, which could make it a useful tool in helping less experienced prostate radiologists perform more accurately, to address the growing shortage of subspecialist expert radiologists^26^.

It is worth noting that the bpMRI AI model achieved better performance when incorporating an automated segmentation of the prostate. This result suggests it can be more efficient and accurate to train a model with known prior anatomical information.

RSIrs-max, initially trained on a small cohort of 46 patients from a single institution, demonstrates strong generalization due to its biophysical basis and alignment with cancer cell morphology^1,2,6,17,27^. Conversely, AI models, are known to benefit from larger and more diverse training datasets. We found that expanding the training data (i.e., moving from Data Split 1 to Data Split 2) improved the AI model performance slightly from (AUC, 0.73 (95% CI: 0.69-0.77)) in Data Split 1 to (AUC, 0.78 (95% CI: 0.73-0.83)) in Data Split 2. These are different datasets, so the AUCs are not directly comparable, but neither PI-RADS nor RSIrs-max saw a similar boost in performance between the two data splits. With larger datasets, combining AI and RSI may even achieve superior performance.

The combination of RSIrs-max or the best deep learning model (Model **6**) with PI-RADS achieved statistically significantly better results than PI-RADS alone. This finding suggests that both RSIrs-max and the deep learning model can serve as complementary tools to PI-RADS for detection of csPCa.

The performance of our models are concordant with other recent studies of AI for prostate MRI that also showed patient-level csPCa detection comparable to PI-RADS^12,28^. For example, in the PI-CAI challenge, an AI model was developed as a single combination of the 5 best performing models (among 293 submitted for the challenge) and performed similarly to radiologists^12^. Of note, beyond imaging data, the PI-CAI model also included age, prostate-specific antigen (PSA) level, prostate volume, and scanner name. Another recent study described two models developed for patient-level detection: one with only imaging data, and another that combined imaging with PSA and PSA density^28^. Performance of AI models is best assessed in external validation datasets. Saha et al. (PI-CAI study) tested their model in data from 1,000 patients from four centers, all using Siemens Healthineers scanners (mainly Siemens Skyra)^12^. Cai et al. tested their model in data from 604 patients from 3 sites of a single academic institution using 2 scanner vendors (Siemens Healthineers and GE Healthcare) and an external dataset^28^. Our Data Split 1 had 664 patients in the testing set from 6 imaging centers and 2 scanner vendors (Siemens Healthineers and GE Healthcare). Data Split 2 had 384 patients in the testing set from 5 imaging centers and the same 2 scanner vendors (Siemens Healthineers and GE Healthcare).

Limitations of our study include those typical for prostate MRI: (1) biopsy is an imperfect gold standard due to possibility of missing csPCa, even when targeting with MRI, and (2) individuals with hip implants were excluded because of known potential to cause severe artifacts on MRI. Larger datasets could facilitate improved AI training, smaller confidence intervals, and increased statistical power to detect differences between models. To our knowledge, though, this is the largest study to date to combine AI and an advanced MRI biomarker for prostate cancer. Finally, we evaluated only one type of deep learning model here; future research will include additional AI architectures.

## Conclusions

Deep learning bpMRI and the RSIrs-max imaging biomarker achieve performance comparable to expert radiologists for detecting csPCa. Combining PI-RADS with RSIrs-max or an AI model outperformed PI-RADS alone, suggesting that both could serve as valuable complements to human expertise. Larger datasets may reveal advantages to integrating RSI in AI.

## Data Availability

All data produced in the present study are available upon reasonable request to the authors.

