## Supplementary Materials for "Deep learning AI and Restriction Spectrum Imaging for patient-level detection of clinically significant prostate cancer on MRI"

**RSI model equation**

The RSI model equation shows the relationship between the DWI signal from the four compartments and the DWI signal intensity at a specific *b*-value. $S\left( b \right)$ represents the measured diffusion-weighted imaging (DWI) signal intensity at a specific *b*-value.

$S\left( b \right) = \sum_{i = 1}^{4} C_{i}e^{-bD_{i}}$	(1)

The signal is computed through the combination of exponential decays, each representing one of four diffusion compartments. $C_{i}$ is the signal contribution of a particular compartment to the overall signal and is determined through model-fitting. $D_{i}$ is the diffusion coefficient that is empirically determined for each of the four compartments $\left( C_{i} \right)$. The detailed parameter for each RSI compartments is shown in supplementary table 1.

**Preprocessing and augmentation of Images**

All 3D-data was first resampled onto the voxel space of the corresponding T2w volume. Then, the resolution of all data was resampled to 0.5 x 0.5 x 3 mm^3^. The 3D data were cropped to 256 x 256 x 32 voxels and normalized to have a zero mean and unit standard deviation. Data augmentation techniques included: random axis mirror flip with a probability of 0.2 for all three dimensions, random intensity scaling in the range (0.9, 1.1) and random intensity shift in the range (-0.1, 0.1).

**Model Training**

The models were trained on 1 NVIDIA GeForce RTX 2080 Ti GPU, with a batch size of 8 for a total of 120 epochs. The learning rate for both models was set to 0.0001. A linear warm-up with a cosine annealing learning rate scheduler was used with 50 warm up epochs.

| **RSI compartment** | **Fixed Diffusion Coefficient (s/mm^2^)** |
| --- | --- |
| Restricted Diffusion $\left( C_{1} \right)$ | $1.1 \times{10}^{-4} \left( D_{1} \right)$ |
| Hindered Diffusion $\left( C_{2} \right)$ | $1.8 \times{10}^{-3} \left( D_{2} \right)$ |
| Free Diffusion $\left( C_{3} \right)$ | $3.6 \times{10}^{-3} \left( D_{3} \right)$ |
| Vascular Flow $\left( C_{4} \right)$ | $0.1220 \left( D_{4} \right)$ |

Supplementary Table 1. RSI compartment and its fixed diffusion coefficient.

| **Institution** | **Scanner models** | **Number of Stations** |
| --- | --- | --- |
| UCSD CTIPM | GE Healthcare Discovery MR750, GE Healthcare Signa Premier | 4 |
| UCSD Health | GE Healthcare Discovery MR750, GE Healthcare Signa Premier | 4 |
| URMC | SIEMENS Magnetom Skyra | 2 |
| MGH | GE Healthcare Signa Premier | 1 |
| UCSF | GE Healthcare Signa Premier | 2 |
| Cambridge | GE Healthcare Discovery MR750 | 1 |
| UTHSCSA | SIEMENS Magnetom Skyra, SIEMENS Magnetom Trio | 3 |
| **Total** | 4 scanner models | 17 stations |

Supplementary Table 2. Scanner models and number of stations for each cohort. UCSD: University of California San Diego. CTIPM: Center for Translational Imaging and Precision Medicine. MGH: Harvard University’s Massachusetts General Hospital. URMC: University of Rochester Medical Center. UTHSCSA: University of Texas Health Sciences Center San Antonio. UCSF: University of California San Francisco. Cambridge: University of Cambridge

| **UCSD CTIPM** | **DWI** | ***T_2_*-weighted** |
| --- | --- | --- |
| Pulse sequence | Diffusion-weighted EPI | Fast Spin Echo (FSE) |
| TR (ms) | 4500 | 7000 |
| TE (ms) | 69 | 100 |
| FOV (mm) | 240 x 120 | 240 x 240 |
| Matrix [resampled dimensions] | 96 x 48 [128 x 128] | 320 x 320 [512 x 512] |
| Slices | 16 | 32 |
| Slice Thickness (mm) | 6 | 3 |
| b-values (s/mm^2^) [number of samples] | 0 [1], 500 [6], 1000 [6], 2000 [12] | N/A |
| Field Strength (T) | 3 | 3 |
| **UCSD Health** | **DWI** | ***T_2_*-weighted** |
| Pulse sequence | Diffusion-weighted EPI | Fast Spin Echo (FSE) |
| TR (ms) | 4000 | 5300 |
| TE (ms) | 69 | 100 |
| FOV (mm) | 240 x 120 | 200 x 200 |
| Matrix [resampled dimensions] | 96 x 48 [256 x 256] | 320 x 320 [512 x 512] |
| Slices | 16 | 32 |
| Slice Thickness (mm) | 6 | 3 |
| b-values (s/mm^2^) [number of samples] | 0 [1], 500 [8], 1000 [8], 2000 [16] | N/A |
| Field Strength (T) | 3 | 3 |
| **URMC** | **DWI** | ***T_2_*-weighted** |
| Pulse sequence | Diffusion-weighted EPI | Fast Spin Echo (FSE) |
| TR (ms) | 3800 | 4800 |
| TE (ms) | 85 | 104 |
| FOV (mm) | 52 x 52 | 180x180 |
| Matrix [resampled dimensions] | 100 x 52 [104 x 200] | 384 x 365 [384 x 384] |
| Slices | 22 | 32 |
| Slice Thickness (mm) | 4 | 3 |
| b-values (s/mm^2^) [number of samples] | 0 [1], 500 [6], 1000 [6], 2000 [6] | N/A |
| Field Strength (T) | 3 | 3 |
| **MGH** | **DWI** | ***T_2_*-weighted** |
| Pulse sequence | Diffusion-weighted EPI | Fast Spin Echo (FSE) |
| TR (ms) | 4500 | 3937 |
| TE (ms) | 59 | 169 |
| FOV (mm) | 240 x 120 | 160 x 160 |
| Matrix [resampled dimensions] | 96 x 48 [128x128] | 360 x 224 [1024 x 1024] |
| Slices | 16 | 40 |
| Slice Thickness (mm) | 6 | 3 |
| b-values (s/mm^2^) [number of samples] | 0 [1], 500 [6], 1000 [6], 2000 [12] | N/A |
| Field Strength (T) | 3 | 3 |
| **Cambridge** | **DWI** | ***T_2_*-weighted** |
| Pulse sequence | Diffusion-weighted EPI | Fast Spin Echo (FSE) |
| TR (ms) | 4500 | 3130 |
| TE (ms) | 68 | 98 |
| FOV (mm) | 220 x 110 | 180 x 180 |
| Matrix [resampled dimensions] | 96 x 48 [256 x 256] | 448 x 256 [512 x 512] |
| Slices | 8 | 30 |
| Slice Thickness (mm) | 4 | 3 |
| b-values (s/mm^2^) [number of samples] | 0 [1], 500 [2], 1000 [2], 2000 [4] | N/A |
| Field Strength (T) | 3 | 3 |
| **UTHSCSA** | **DWI** | ***T_2_*-weighted** |
| Pulse sequence | Diffusion-weighted EPI | Fast Spin Echo (FSE) |
| TR (ms) | 6300 | 4710 |
| TE (ms) | 105 | 100 |
| FOV (mm) | 52 x 100 | 180 x 180 |
| Matrix [resampled dimensions] | 52 x 100 [104 x 200] | 240 x 320 [320 x 320] |
| Slices | 22 | 30 |
| Slice Thickness (mm) | 4 | 3 |
| b-values (s/mm^2^) [number of samples] | 0 [1], 500 [6], 1000 [18], 2000 [42] | N/A |
| Field Strength (T) | 3 | 3 |
| **UCSF** | **DWI** | ***T_2_*-weighted** |
| Pulse sequence | Diffusion-weighted EPI | Fast Spin Echo (FSE) |
| TR (ms) | 4500 | 2964 |
| TE (ms) | 73 | 150 |
| FOV (mm) | 200 x 200 | 220 x 220 |
| Matrix [resampled dimensions] | 256 x 256 [256 x 256] | 512 x 512 [320 x 320] |
| Slices | 35 | 36 |
| Slice Thickness (mm) | 3 | 3 |
| b-values (s/mm^2^) [number of samples] | 0 [5], 100 [6], 800 [12], 1400 [12], 2500 [18] | N/A |
| Field Strength (T) | 3 | 3 |

Supplementary Table 3. MRI acquisition parameters for each cohort. TR: repetition time. TE: echo time. FOV: field-of-view. FSE: fast spin echo. EPI: echo-planar imaging. DWI: diffusion-weighted imaging. UCSD: University of California San Diego. CTIPM: Center for Translational Imaging and Precision Medicine. MGH: Harvard University’s Massachusetts General Hospital. URMC: University of Rochester Medical Center. UTHSCSA: University of Texas Health Sciences Center San Antonio. UCSF: University of California San Francisco. Cambridge: University of Cambridge
